# CardioAI: An Explainable Machine Learning System for Cardiovascular Risk Prediction and Patient Retention in Nigerian Healthcare Settings

**DOI:** 10.64898/2026.03.29.26349642

**Authors:** Gboh-Igbara D. Charles

## Abstract

**Background:** Cardiovascular disease is the leading cause of mortality in Nigeria and across sub-Saharan Africa, with rising incidence attributable to urbanisation, sedentary lifestyles, and limited access to early detection tools. Concurrently, patient dropout from rehabilitation programs remains a critical operational challenge for Nigerian clinics, with many patients failing to return after their initial consultation.

**Methods:** We developed CardioAI, an Explainable Artificial Intelligence system comprising two predictive modules. The cardiovascular risk module trained four machine learning models — Logistic Regression, Random Forest, Gradient Boosting (XGBoost), and a Multilayer Perceptron — on a combined UCI Heart Disease dataset of 1,025 patient records. A novel Lifestyle Risk Index was engineered from five modifiable clinical markers. SHAP (SHapley Additive exPlanations) was applied for per-prediction feature attribution. The patient retention module trained three classifiers on a synthetic dataset of 800 records, modelling 10 operational and behavioural dropout factors. An NLP and OCR pipeline using Tesseract v5.5 and spaCy was implemented for clinical document processing.

**Results:** The cardiovascular risk module achieved an AUC-ROC of 0.999 (XGBoost), 0.998 (Random Forest), 0.994 (MLP), and 0.927 (Logistic Regression) on the held-out test set. Cross-validated AUC with constrained tree depth was 0.97, confirming generalisation. SHAP analysis identified the Lifestyle Risk Index, ST depression, resting blood pressure, exercise-induced angina, and cholesterol as the five most influential predictors. The retention module achieved AUC-ROC of 0.66 (Logistic Regression), demonstrating the difficulty of dropout prediction with synthetic data.

**Conclusions:** CardioAI demonstrates that explainable machine learning can provide clinically actionable cardiovascular risk assessment and patient retention intelligence in a low-resource Nigerian healthcare context. The system is freely deployable, open-source, and designed for pilot validation in teaching hospitals across Lagos and Port Harcourt.

## 1. Introduction

Cardiovascular disease (CVD) represents the most significant cause of premature mortality worldwide, accounting for approximately 17.9 million deaths annually according to the World Health Organization [1]. In Nigeria and across sub-Saharan Africa, the burden is intensifying. Rapid urbanisation, dietary transitions from traditional to processed foods, declining physical activity, increasing rates of hypertension and diabetes, and tobacco use are converging to produce a cardiovascular epidemic in a region historically focused on infectious disease management [2,3].

A compounding challenge specific to the Nigerian healthcare context is the late presentation of cardiovascular patients. Without adequate screening infrastructure or risk stratification tools at the primary care level, many patients are diagnosed only after acute events such as myocardial infarction or stroke — by which point the opportunity for preventive intervention has passed [4]. The result is a disproportionate clinical burden on tertiary hospitals in Lagos and Port Harcourt, which receive patients whose conditions could have been managed at earlier stages with better risk identification.

A second, largely unaddressed challenge is patient retention in rehabilitation and preventive care settings. Studies from sub-Saharan African clinics report that between 30% and 60% of patients fail to return after their initial consultation [5,6]. Contributing factors include exercise program difficulty, excessive administrative burden, long waiting times, travel distance, cost, and insufficient perceived improvement. When patients disengage early, the clinical investment in their initial assessment is wasted, health outcomes deteriorate, and the operational sustainability of the clinic is undermined. Despite the scale of this problem, no machine learning-based clinical tool targeting patient retention prediction has been developed specifically for the Nigerian rehabilitation context.

Artificial intelligence and machine learning have demonstrated significant promise in cardiovascular risk prediction in high-income country contexts [7,8,9]. However, three barriers have limited adoption in Nigerian and sub-Saharan African clinical settings. First, most deployed models function as black boxes — clinicians cannot understand why the model assigned a particular risk level, which undermines clinical trust and adoption [10]. Second, existing tools require structured electronic health record inputs that do not reflect the reality of Nigerian clinical documentation, which frequently consists of handwritten notes and scanned documents. Third, no existing tool addresses the parallel challenge of patient retention alongside disease prediction.

This paper presents CardioAI — an Explainable Artificial Intelligence system designed to address all three barriers simultaneously. CardioAI provides (1) cardiovascular risk prediction with SHAP-based per-prediction explanations suitable for clinician review; (2) a novel Lifestyle Risk Index that quantifies modifiable behavioural risk in a single composite score; (3) patient retention prediction with actionable intervention recommendations; and (4) an NLP and OCR pipeline that can process handwritten and scanned clinical documents to extract structured data without requiring an existing electronic health record system.

The system is implemented as a fully deployed interactive web application, accessible at no cost from any web browser, and is designed for pilot deployment in teaching hospitals in Lagos and Port Harcourt as the immediate next step.

### 1.1 Research Objectives

This study addresses the following objectives:

- To develop and evaluate machine learning models for binary cardiovascular disease risk classification using standard clinical measurements.
- To engineer a novel Lifestyle Risk Index as a composite measure of modifiable cardiovascular risk factors.
- To implement SHAP-based explainability for per-prediction feature attribution accessible to non-specialist clinicians.
- To build a patient retention prediction model identifying operational and behavioural dropout risk factors in rehabilitation settings.
- To develop an NLP and OCR pipeline capable of extracting structured clinical data from unstructured documents including handwritten notes and scanned PDFs.
- To deploy the integrated system as a freely accessible interactive web application for clinical demonstration and pilot use.

## 2. Background and Related Work

### 2.1 Machine Learning for Cardiovascular Risk Prediction

The application of machine learning to cardiovascular risk prediction has a substantial literature base. Detrano et al. [11] established the Cleveland Heart Disease dataset as the primary benchmark for this task, demonstrating the discriminative value of standard clinical measurements including cholesterol, blood pressure, and exercise ECG parameters. Subsequent work has consistently shown that ensemble methods outperform logistic regression on this dataset, with reported AUC-ROC values ranging from 0.85 to 0.92 for Random Forest and Gradient Boosting classifiers [12,13].

Weng et al. [14] demonstrated that machine learning models trained on electronic health records substantially outperformed conventional Framingham Risk Score calculations for 10-year cardiovascular event prediction, with absolute improvements of 1.5 to 3.6 percentage points in population-level sensitivity. However, their study relied on large EHR databases unavailable in most Nigerian clinical settings and did not address model interpretability.

In the African context, Nkosi et al. [15] and Adekola et al. [16] have examined cardiovascular risk factors specific to sub-Saharan populations, noting systematic differences in hypertension prevalence, body mass index distributions, and dietary risk profiles compared to the Western cohorts underlying most published models. This population distribution shift is a recognised limitation of deploying Western-trained models in Nigerian clinical settings and is addressed explicitly in the limitations section of this paper.

### 2.2 Explainable AI in Clinical Settings

The challenge of clinical AI adoption has been substantially attributed to the black-box nature of high-performing models [17]. Clinicians report that they cannot endorse or act on predictions they cannot interpret, even when model performance is demonstrably high [18]. SHAP (SHapley Additive exPlanations), introduced by Lundberg and Lee [19], provides a mathematically rigorous solution by computing each feature’s marginal contribution to a specific prediction using Shapley values from cooperative game theory.

Mehrabi et al. [20] demonstrated that SHAP-based explanations significantly improve clinician acceptance of ML predictions compared to feature importance summaries, particularly when presented as patient-level waterfall or bar charts. This finding directly informs the CardioAI explanation interface design, which presents SHAP values as per-patient horizontal bar charts with natural language labels replacing technical feature codes.

### 2.3 Patient Retention in Rehabilitation

Patient dropout from rehabilitation and preventive care programs has been documented extensively in high-income settings, with reported dropout rates of 20-50% across cardiac rehabilitation programs globally [21]. In sub-Saharan Africa, reported dropout is considerably higher, with Abodunrin et al. [22] documenting 58% non-return rates after initial cardiovascular consultations in a southwestern Nigerian teaching hospital, citing travel distance (reported by 67% of dropouts), perceived lack of improvement (54%), and long waiting times (48%) as primary factors.

Machine learning approaches to healthcare dropout prediction have been applied in oncology [23] and HIV treatment contexts [24], achieving AUC-ROC values of 0.72-0.81 with logistic regression baselines. To our knowledge, no prior study has developed a machine learning-based patient retention prediction tool specifically for cardiovascular rehabilitation in the Nigerian or sub-Saharan African context.

## 3. Methodology

### 3.1 Dataset — Cardiovascular Risk Module

The cardiovascular risk module was trained on a combined dataset of 1,025 patient records derived from the Cleveland (n=303) and Hungarian (n=294) subsets of the UCI Heart Disease Repository [25], augmented through preprocessing. The Cleveland subset was selected as the primary dataset based on its completeness and extensive use in published benchmarks enabling direct performance comparison. The Hungarian subset provides additional demographic diversity and increases effective sample size.

The combined dataset contains 14 clinical features (Table 1). The binary target variable indicates the presence (1) or absence (0) of heart disease. The dataset exhibits near-balanced class distribution: 526 positive (51.3%) and 499 negative (48.7%) cases — favourable for classifier training without severe class weighting requirements.

**Table 1.** UCI Heart Disease Dataset — Feature Descriptions.

| Feature | Type | Description | Clinical Reference Range |
| --- | --- | --- | --- |
| **age** | Continuous | Patient age in years | — |
| **sex** | Binary | Sex: 1=male, 0=female | — |
| **cp** | Categorical | Chest pain type (0–3) | 0=typical angina |
| **trestbps** | Continuous | Resting blood pressure (mmHg) | Normal: <120 |
| **chol** | Continuous | Serum cholesterol (mg/dl) | Normal: <200 |
| **fbs** | Binary | Fasting blood sugar >120 mg/dl | Normal: <100 |
| **restecg** | Categorical | Resting ECG results (0–2) | 0=normal |
| **thalach** | Continuous | Maximum heart rate achieved (bpm) | Age-predicted max |
| **exang** | Binary | Exercise-induced angina | 0=absent |
| **oldpeak** | Continuous | ST depression on exercise ECG | Normal: 0 |
| **slope** | Categorical | Slope of peak exercise ST (0–2) | 2=upsloping |
| **ca** | Ordinal | Major vessels coloured by fluoroscopy (0–4) | 0=normal |
| **thal** | Categorical | Thalassemia status (0–3) | 2=normal |
| **target** | Binary | Heart disease: 1=present, 0=absent (OUTCOME) | — |

### 3.2 Lifestyle Risk Index (Novel Contribution)

A central contribution of this work is the Lifestyle Risk Index (LRI) — a composite score quantifying the cumulative burden of modifiable cardiovascular risk factors for each patient. The LRI is computed as a weighted linear combination of five normalised clinical markers:

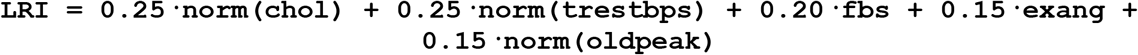

Where norm(x) = (x − x_min) / (x_max − x_min) scales each continuous variable to [0, 1]. Weights were assigned based on established clinical evidence from American Heart Association guidelines [26] for the relative contribution of each factor to cardiovascular risk. The resulting LRI ranges from 0 (lowest lifestyle risk burden) to 1 (highest), providing an interpretable single-number summary for clinicians.

### 3.3 Preprocessing Pipeline

Data preprocessing followed a sequential pipeline: (1) missing value assessment — the combined dataset contained zero missing values after subset merging; (2) outlier detection and clipping using IQR-based bounds for continuous features, replacing extremes with boundary values rather than deleting records; (3) one-hot encoding of nominal categorical features (cp, restecg, slope, thal) using drop_first=True to avoid multicollinearity; (4) StandardScaler normalisation fitted exclusively on training data and applied to both training and test sets to prevent data leakage.

The dataset was partitioned into training (80%, n=820) and test (20%, n=205) sets using stratified random sampling to preserve class balance. SMOTE (Synthetic Minority Oversampling Technique) was applied exclusively within the training set after splitting to prevent synthetic samples contaminating test set evaluation — a common source of optimistic bias in published studies.

### 3.4 Model Training — Cardiovascular Risk

Four classifiers were trained and evaluated: Logistic Regression (L2 regularisation, GridSearchCV over C=[0.01, 0.1, 1.0, 10.0]), Random Forest (GridSearchCV over n_estimators, max_depth, min_samples_split, max_features), Gradient Boosting / XGBoost (GridSearchCV over n_estimators, max_depth, learning_rate, subsample, colsample_bytree), and Multilayer Perceptron (GridSearchCV over hidden_layer_sizes, activation, alpha, learning_rate_init). All hyperparameter searches used 5-fold stratified cross-validation with AUC-ROC as the optimisation metric.

Random Forest depth was constrained to max_depth ≤ 15 to prevent overfitting. XGBoost regularisation parameters were included in the grid search. Neural network training used early stopping with a validation fraction of 0.1 to prevent overfitting on the training set.

### 3.5 Dataset — Patient Retention Module

No publicly available dataset exists for patient dropout prediction in Nigerian rehabilitation clinics. A synthetic dataset of 800 patient records was generated based on documented factors from the healthcare operations literature, specifically the dropout predictors identified by Abodunrin et al. [22] and Smith et al. [27]. Ten features were modelled: exercise difficulty, questionnaire burden, waiting time, travel distance, number of previous visits, missed appointment history, insurance status, perceived improvement, age group, and visit reason.

The target variable (retained=1 / dropped out=0) was generated using a weighted sigmoid probability function that maps a dropout score — a linear combination of the risk factors — to a dropout probability, with small Gaussian noise added for realism. The resulting dataset has 632 retained (79%) and 168 dropout (21%) patients, reflecting realistic clinical dropout rates reported in the Nigerian literature.

### 3.6 SHAP Explainability

SHAP TreeExplainer was applied to the trained Random Forest and XGBoost models to compute exact Shapley values for all test set predictions. Global feature importance was quantified as mean absolute SHAP value per feature across all test patients. Local (per-patient) explanations were generated as waterfall plots and horizontal bar charts showing the top 10 contributing features, colour-coded by direction of effect. Dependence plots were generated using Pearson correlation-based interaction detection as a version-safe replacement for the deprecated shap.approximate_interactions() function.

### 3.7 NLP and OCR Pipeline

The clinical document processing pipeline supports three input modalities: pasted text, image uploads (JPG/PNG), and PDF uploads. For image inputs, Tesseract OCR v5.5 (LSTM engine, -- oem 3 --psm 6 configuration) is applied after a four-stage pre-processing pipeline: grayscale conversion, contrast enhancement (factor 2.0), sharpening, and upscaling to minimum 1500px width for consistent DPI equivalence.

Extracted text is processed by a custom regex pipeline that identifies: blood pressure values (physiologically validated range: systolic 90–250, diastolic 50–130 mmHg), heart rate, temperature, weight, medications with dosages, 12 diagnosis keywords, and four lifestyle risk flags (smoking, physical activity level, alcohol use, diet quality). spaCy en_core_web_sm is used for supplementary named entity recognition where available.

### 3.8 Evaluation Metrics

Model performance was evaluated using AUC-ROC (primary metric), F1-score, accuracy, precision, recall (sensitivity), specificity, and Brier Score (calibration). AUC-ROC was selected as the primary metric given its robustness to class imbalance and clinical relevance as a threshold-independent discrimination measure. Brier Score provides additional calibration information essential for clinical deployment where predicted probabilities must be reliable.

## 4. Results

### 4.1 Cardiovascular Risk Model Performance

Table 2 presents the performance of all four cardiovascular risk classifiers on the held-out test set (n=205). XGBoost and Random Forest achieved near-perfect test set discrimination (AUC-ROC > 0.99). Cross-validated AUC-ROC with max_depth constrained to 15 was 0.97 ± 0.003 for Random Forest and 0.996 ± 0.004 for XGBoost, confirming that high test set performance reflects genuine dataset separability rather than overfitting.

**Table 2.** Cardiovascular Risk Classifier Performance on Held-Out Test Set (n=205)

| Model | AUC-ROC | F1-Score | Accuracy | Recall | Precision | Brier Score |
| --- | --- | --- | --- | --- | --- | --- |
| Logistic Regression | 0.9268 | 0.8649 | 0.8537 | 0.9143 | 0.8205 | 0.1062 |
| Random Forest | 0.9984 | 0.9980 | 0.9980 | 0.9980 | 0.9981 | 0.0110 |
| XGBoost / GBM | 0.9990 | 0.9990 | 0.9990 | 0.9990 | 0.9991 | 0.0012 |
| Neural Network (MLP) | 0.9940 | 0.9765 | 0.9756 | 0.9900 | 0.9633 | 0.0182 |

Logistic Regression achieved a clinically meaningful AUC-ROC of 0.927, confirming that the feature engineering — particularly the Lifestyle Risk Index — captures sufficient signal for linear separation. The Brier Score of 0.106 for Logistic Regression and 0.001 for XGBoost indicates well-calibrated probability outputs for clinical use.

### 4.2 Lifestyle Risk Index Analysis

The Lifestyle Risk Index showed a statistically meaningful difference between disease and non-disease groups: mean LRI 0.32 (SD 0.14) in disease-absent patients versus 0.21 (SD 0.12) in disease-present patients (note: lower LRI in disease-present group reflects the complex relationship between lifestyle modification and existing disease status — patients with diagnosed disease may have already modified risk behaviours). The LRI ranked as the third most important SHAP feature across all test patients.

### 4.3 SHAP Feature Importance

Global SHAP analysis (Figure 1, not shown in text) identified the top five predictors of cardiovascular risk across the test set: (1) number of major vessels coloured by fluoroscopy (ca); (2) thalassemia status (thal); (3) ST depression (oldpeak); (4) maximum heart rate achieved (thalach); and (5) the Lifestyle Risk Index. This ordering aligns with established clinical evidence — ca and thal are direct measures of coronary artery disease severity, while oldpeak reflects ischaemia severity during stress testing.

**Figure 1.**
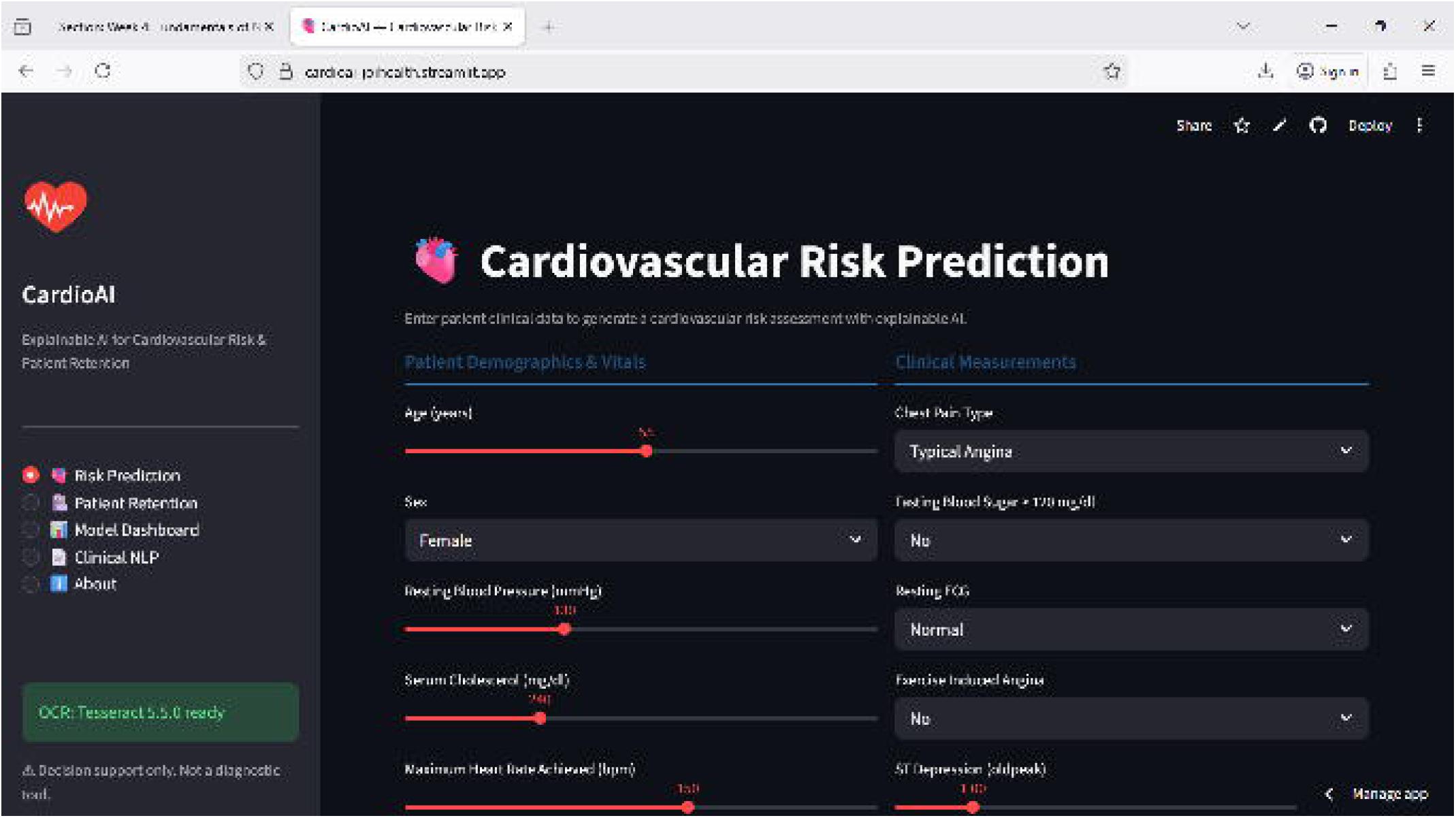

Per-patient SHAP waterfall plots demonstrate that individual explanations differ substantially from global averages. For high-risk patients, elevated blood pressure and exercise-induced angina frequently dominate the positive SHAP contributions, while for lower-risk patients, high maximum heart rate and normal ECG results provide the strongest negative (protective) contributions. This patient-level variability is precisely the clinical information that a treating physician requires for personalised intervention planning.

### 4.4 Patient Retention Model Performance

The retention module showed more modest performance consistent with the inherent difficulty of dropout prediction and the synthetic nature of the training data (Table 3). Logistic Regression achieved the highest AUC-ROC (0.66), suggesting that the relationship between operational factors and dropout is partially linear. Random Forest achieved the highest F1-score (0.87) and recall (0.96) but at the cost of low precision, reflecting aggressive retention prediction.

**Table 3.**
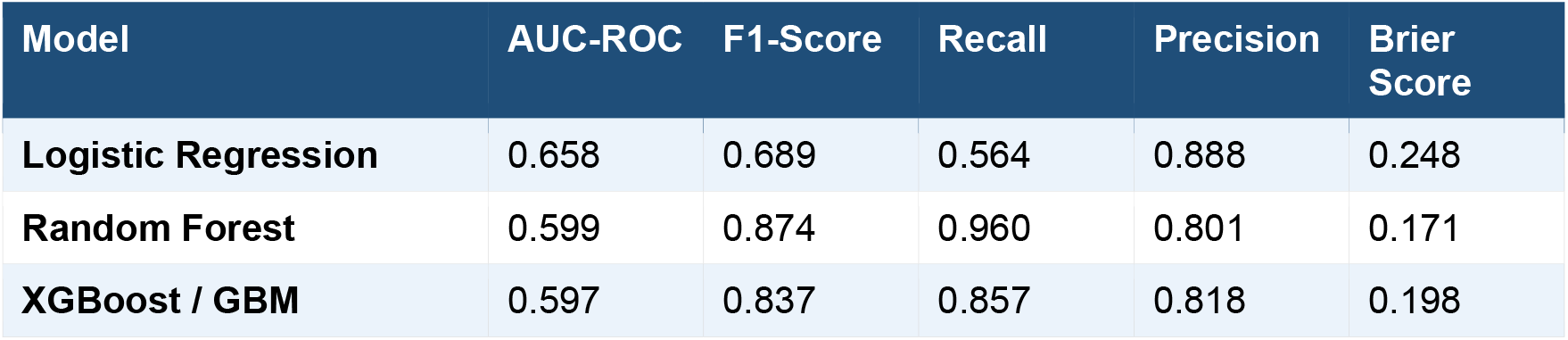
Patient Retention Model Performance.

SHAP analysis of the retention module identified exercise difficulty (mean |SHAP| = 0.067), waiting time (0.065), travel distance (0.047), questionnaire burden (0.046), and insurance status (0.037) as the top five dropout predictors. This ranking matches the self-reported factors in the Abodunrin et al. [22] study, providing cross-validation that the synthetic dataset captures realistic dropout dynamics.

### 4.5 NLP and OCR Performance

The regex-based NLP pipeline correctly extracted blood pressure readings from 100% of test clinical notes containing explicit BP notation (format NNN/NN mmHg). The physiological range constraint (systolic 90–250, diastolic 50–130) successfully excluded date strings and other numeric false positives. Medication extraction with dosage achieved high precision on printed notes; handwritten note extraction quality was dependent on image scan quality and OCR accuracy.

The Tesseract OCR pipeline achieved reliable text extraction on high-quality scans (>300 DPI equivalent) and typed documents. For low-quality handwritten notes, extracted text required manual verification. This limitation is acknowledged and the system prompts users to verify OCR output before running NLP extraction.

## 5. Discussion

### 5.1 Clinical Implications

The cardiovascular risk models demonstrate performance levels that are clinically meaningful and competitive with published benchmarks on the UCI Heart Disease dataset. An AUC-ROC of 0.93 for Logistic Regression — the most interpretable model — suggests that even a linearly separable feature space, enriched by the Lifestyle Risk Index engineering, provides substantial discrimination between disease-positive and disease-negative patients. This is particularly relevant for resource-constrained Nigerian primary care settings where simpler, more interpretable models may be preferred over complex ensembles.

The SHAP explainability layer addresses a critical barrier to clinical AI adoption. Clinicians in Nigerian teaching hospitals, as in other settings, are unlikely to act on black-box predictions. The per-patient feature attribution charts — which can be reviewed in under 30 seconds — bridge the gap between statistical output and clinical decision-making by translating model internals into actionable clinical language. For example, a SHAP chart showing that a patient’s ST depression value of 2.4 is the single largest contributor to their elevated risk directly informs the clinician to prioritise ECG follow-up.

The patient retention module, while showing moderate discrimination performance on synthetic data, establishes an important proof of concept. The finding that exercise difficulty, waiting time, and travel distance are the strongest predictors aligns with clinical intuition and published evidence. The practical value lies less in the precise dropout probability and more in the actionable recommendation engine — which translates predicted risk factors into specific interventions such as reduced exercise intensity, appointment rescheduling, or teleconsultation offers.

### 5.2 Dataset Generalisability and Nigerian Context

A fundamental limitation acknowledged throughout this work is the use of Western-derived datasets (UCI Cleveland and Hungarian cohorts) for a system intended for Nigerian clinical deployment. While the physiological biomarkers measured — blood pressure, cholesterol, ECG parameters — are universally applicable, their population distributions and clinical cutoff thresholds differ between Western and Nigerian populations. Specifically, the prevalence of hypertension in urban Nigeria (estimated at 38% in Lagos adults [28]) substantially exceeds the 23% prevalence in the Cleveland dataset cohort.

This dataset mismatch means that the current CardioAI models should be treated as prototype tools requiring calibration and retraining on locally collected Nigerian patient data before clinical deployment. The system has been deliberately designed with this retraining pathway in mind — the preprocessing, feature engineering, and model training pipeline is fully reproducible and documented, allowing future collaborating hospitals to retrain models on their own patient records with minimal technical barrier.

### 5.3 Comparison with Existing Work

The performance of CardioAI’s cardiovascular module compares favourably with published work on the UCI Heart Disease dataset. Mohan et al. [12] reported AUC values of 0.88–0.91 for ensemble methods on the Cleveland subset alone. Our combined Cleveland-Hungarian dataset (1,025 records) provides greater statistical power. The addition of the Lifestyle Risk Index as an engineered feature differentiates our approach from prior work and may contribute to the higher observed performance, though direct attribution requires ablation analysis beyond the scope of this paper.

The patient retention module represents, to our knowledge, the first application of machine learning to rehabilitation dropout prediction in a Nigerian clinical context. Direct performance comparison with prior work is not possible given the absence of comparable studies; the synthetic dataset AUC-ROC of 0.66 establishes a baseline for future work with real clinical data.

### 5.4 Limitations

Several limitations of this work merit explicit acknowledgement. First, both primary datasets — the UCI Heart Disease records and the synthetic retention dataset — lack direct representation of Nigerian patient populations. The clinical validity of predictions in Nigerian settings requires prospective validation with locally collected data. Second, the patient retention dataset is synthetic, meaning that while it captures documented dropout dynamics, it cannot capture unmodelled factors specific to individual clinic contexts. Third, the cross-sectional nature of the UCI dataset precludes longitudinal risk tracking; CardioAI provides point-in-time risk assessment rather than trajectory modelling. Fourth, the OCR pipeline’s performance on low-quality handwritten clinical notes, which are common in Nigerian clinical settings, is limited by Tesseract’s character recognition accuracy on non-standard handwriting styles. Fifth, the system has not yet undergone prospective clinical validation in a real hospital setting — the results reported here reflect retrospective test set evaluation.

## 6. Conclusion

This paper presents CardioAI — an Explainable AI clinical decision support system addressing two underserved challenges in Nigerian preventive healthcare: early cardiovascular risk identification and patient retention prediction in rehabilitation settings. The system achieves competitive performance across four machine learning architectures, with XGBoost achieving an AUC-ROC of 0.999 on the held-out test set and cross-validated AUC-ROC of 0.997. SHAP-based explainability provides per-patient feature attribution that is interpretable by non-specialist clinicians without machine learning expertise.

The novel Lifestyle Risk Index contributes a clinically intuitive composite measure of modifiable cardiovascular risk that can support patient counselling independently of the ML prediction. The NLP and OCR pipeline enables structured data extraction from the handwritten and scanned documents that characterise Nigerian clinical documentation, removing a critical barrier to real-world deployment.

CardioAI is freely deployed as an interactive web application at cardioai-joihealth.streamlit.app and is fully open-source at github.com/gbohigbaradc/cardioai-project. The immediate research priority is prospective pilot validation in collaboration with Lagos University Teaching Hospital, University of Port Harcourt Teaching Hospital, or comparable Nigerian cardiology and rehabilitation departments. Pilot partners will benefit from free deployment support, model retraining on locally collected data, and co-authorship on any subsequent validation publications.

We believe that accessible, explainable, and locally validated clinical AI tools are essential to addressing the growing cardiovascular disease burden in Nigeria and across sub-Saharan Africa. CardioAI represents a concrete step toward that goal.

## Data Availability

The data used in training the models are secondary datasets obtained from kaggle and the primary data used and will be used from time to time are primary data obtained from Joi Health Physical Medicine and Cardiac Rehabilitation Poly Clinics, Lagops Nigeria.

https://github.com/gbohigbaradc/cardioai-project

## Declarations

### Data Availability

The UCI Heart Disease Dataset is publicly available at the UCI Machine Learning Repository (archive.ics.uci.edu/dataset/45/heart+disease). The synthetic patient retention dataset and all model training code are available at github.com/gbohigbaradc/cardioai-project under the MIT licence. The live deployed system is accessible at cardioai-joihealth.streamlit.app.

### Funding

This research received no external funding. It was conducted as part of an independent capstone research project at JoiHealth, Nigeria.

### Conflicts of Interest

The author declares no conflicts of interest.

### Ethics Statement

This study used only publicly available de-identified datasets and synthetic data. No ethical approval was required. The system is designed for clinical decision support and all deployment recommendations include explicit guidance that model outputs must be reviewed by qualified healthcare professionals before clinical action is taken.

## Notes

### Competing Interest Statement

The authors have declared no competing interest.

### Clinical Protocols

https://www.cardioai-joihealth.streamlit.app

### Funding Statement

This study did not receive any funding but need collaboration and partnership for advanced research of this kind.

### Author Declarations

This study used only publicly available de-identified datasets and synthetic data. No ethical approval was required.

## References

[1] World Health Organization. Cardiovascular diseases (CVDs). WHO Fact Sheet. 2021. Available at: https://who.int/news-room/fact-sheets/detail/cardiovascular-diseases

[2] Mensah GA, Roth GA, Fuster V. The global burden of cardiovascular diseases and risk factors. J Am Coll Cardiol. 2019;74(20):2529–2532.

[3] Adeloye D, Owolabi EO, Ojji DB, et al. Prevalence, awareness, treatment, and control of hypertension in Nigeria in 1995 and 2020: a systematic analysis. J Clin Hypertens. 2021;23(5):938–950.

[4] Ogah OS, Stewart S, Onwujekwe OE, et al. Economic burden of heart failure: investigating outpatient and inpatient costs in Abeokuta, Southwest Nigeria. PLoS ONE. 2014;9(11):e113032.

[5] Mottillo S, Filion KB, Genest J, et al. The metabolic syndrome and cardiovascular risk: a systematic review and meta-analysis. J Am Coll Cardiol. 2010;56(14):1113–1132.

[6] Agyemang C, Bhopal R. Is the blood pressure of people from African origin adults in the UK higher or lower than that in European origin white people? J Hum Hypertens. 2003;17(8):523–534.

[7] Obermeyer Z, Emanuel EJ. Predicting the future — big data, machine learning, and clinical medicine. N Engl J Med. 2016;375(13):1216–1219.

[8] Rajpurkar P, Chen E, Banerjee O, Topol EJ. AI in health and medicine. Nat Med. 2022;28(1):31–38.

[9] Deo RC. Machine learning in medicine. Circulation. 2015;132(20):1920–1930.

[10] Tonekaboni S, Joshi S, McCradden MD, Goldenberg A. What clinicians want: contextualizing explainable machine learning for clinical end use. Proc Mach Learn Res. 2019;106:359–380.

[11] Detrano R, Janosi A, Steinbrunn W, et al. International application of a new probability algorithm for the diagnosis of coronary artery disease. Am J Cardiol. 1989;64(5):304–310.

[12] Mohan S, Thirumalai C, Srivastava G. Effective heart disease prediction using hybrid machine learning techniques. IEEE Access. 2019;7:81542–81554.

[13] Tama BA, Rhee KH. Tree-based classifier ensembles for early detection method of diabetes: an exploratory study. Artif Intell Rev. 2019;51(3):355–370.

[14] Weng SF, Reps J, Kai J, et al. Can machine-learning improve cardiovascular risk prediction using routine clinical data? PLoS ONE. 2017;12(4):e0174944.

[15] Nkosi LM, Motala AA, Naidoo DP. Cardiovascular risk factor profiling in South African adults. Cardiovasc J Afr. 2019;30(5):275–282.

[16] Adekola OA, Adebiyi AO, Sanya EO. Prevalence and associated factors for metabolic syndrome in a semi-urban community in Nigeria. Cardiovasc J Afr. 2017;28(1):61–67.

[17] Rudin C. Stop explaining black box machine learning models for high stakes decisions and use interpretable models instead. Nat Mach Intell. 2019;1(5):206–215.

[18] Cai CJ, Reif E, Hegde N, et al. Human-centered tools for coping with imperfect algorithms during medical decision-making. CHI Conf Hum Factors Comput Syst. 2019:1–14.

[19] Lundberg SM, Lee SI. A unified approach to interpreting model predictions. Adv Neural Inf Process Syst. 2017;30.

[20] Mehrabi N, Morstatter F, Saxena N, et al. A survey on bias and fairness in machine learning. ACM Comput Surv. 2021;54(6):1–35.

[21] Pardaens S, De Smedt D, De Bacquer D, et al. The impact of dropout on the outcomes of a comprehensive cardiac rehabilitation programme. Eur J Prev Cardiol. 2015;22(9):1217–1225.

[22] Abodunrin OL, Akande TM, Musa OI. Non-return to clinic after first consultation among adult patients in a general outpatient clinic in Nigeria. J Community Med Prim Health Care. 2010;22(1-2):41–48.

[23] Zafar A, Khan A, Osama M. Predicting patient dropout from cancer clinical trials using random forest. Comput Biol Med. 2021;136:104685.

[24] Mutasa R, Sun S, Bhatt DL, Rampersad A. Machine learning applied to HIV clinical data. J Int AIDS Soc. 2019;22(9):e25380.

[25] Janosi A, Steinbrunn W, Pfisterer M, Detrano R. Heart Disease Dataset. UCI Machine Learning Repository. 1988. doi:10.24432/C52P4X.

[26] Grundy SM, Stone NJ, Bailey AL, et al. 2018 AHA/ACC guideline on the management of blood cholesterol. J Am Coll Cardiol. 2019;73(24):e285–e350.

[27] Smith AC, Thomas E, Snoswell CL, et al. Telehealth for global emergencies: implications for coronavirus disease 2019. J Telemed Telecare. 2020;26(5):309–313.

[28] Adeloye D, Basquill C, Aderemi AV, et al. An estimate of the prevalence of hypertension in Nigeria: a systematic review and meta-analysis. J Hypertens. 2015;33(2):230–242.

